# Modeling the impact of case finding for tuberculosis: The role of infection dynamics

**DOI:** 10.1101/2025.04.15.25325877

**Authors:** Theresa S Ryckman, Sourya Shrestha, Anthony T Fojo, Parastu Kasaie, David W Dowdy, Emily A Kendall

## Abstract

**Background:** Our understanding of *Mycobacterium tuberculosis (Mtb)* natural history and infection dynamics is evolving, including recognition that many individuals previously infected with *Mtb* may clear their infections or experience substantially reduced progression risks with time since infection. Such dynamics suggest that recent transmission is more important in driving TB incidence in high-burden settings than previously estimated; thus, the impact of interventions to reduce transmission (e.g., community-based active case-finding) may also be greater than previously thought.

**Methods:** We constructed two models of *Mtb* transmission that differed only in that one model included a clearance mechanism while the other did not. We then calibrated these models independently to the same set of epidemiological data representative of a high-TB-burden setting (India). Finally, we used the calibrated models to project the impact of illustrative biennial active case-finding campaigns (achieving 75% population coverage with 65% screening sensitivity).

**Results:** The model that included a clearance mechanism projected a greater impact of case-finding on the incidence of TB disease: 45% [95% uncertainty interval 28-57%] reduction versus no intervention after 10 years, versus 11% [6-18%] in the model without a clearance mechanism. The estimated annual risk of *Mtb* infection and prevalence of recent infection were both substantially higher in the model that allowed for *Mtb* clearance, despite being fit to the same data.

**Conclusions:** Models that allow for *Mtb* clearance are supported by biological and epidemiological evidence and project greater impact from active case-finding than models that do not include these dynamics.

## Background

Interventions to reduce the global burden of tuberculosis (TB; 10.8 million new cases and 1.3 million deaths in 2023) are urgently needed.^1^ In prioritizing between interventions to find, treat, or prevent TB, it is important to understand that underlying dynamics of infection and progression affect the relative impact of these interventions. Interventions that detect people with undiagnosed TB and link them to treatment (such as community-based “active case-finding”) are likely to have greater impact in epidemics where most TB disease is caused by recent transmission events. If most TB incidence instead arises from reactivation of *Mycobacterium tuberculosis* (*Mtb)* infections that were transmitted in the remote past, then finding and treating people who have TB disease now is unlikely to have major effects on subsequent TB incidence, because TB will continue to develop from within the current pool of remote infections.

Community-based active case-finding programs are globally recommended and are being increasingly prioritized in high-burden settings.^2–5^ Empirical evidence on the impact of active case-finding has been mixed and is challenging to obtain, owing to the need for very large sample sizes to demonstrate effects at a population level.^6^ At least one case-finding study, the ACT3 randomized trial in Vietnam, detected a large and significant reduction in prevalence.^7^ Mathematical models, which are often used to bridge the gap between empirical evidence and policy decisions, have yielded varied estimates of the long-term impact of case-finding. Several modeling studies have projected that case-finding will have only limited and temporary effects on TB incidence, ^8–11^ while others predict greater reductions.^12^

Model-driven estimates of the impact of case-finding may depend on how these models represent underlying *Mtb* infection dynamics – and our understanding of these dynamics is evolving. It is increasingly understood that *Mtb* infection is more complex and dynamic than the conventional concept of a state of lifelong “latency”, and that immunoreactivity to *Mtb* antigens may only weakly reflect current infection.^13–16^ Specifically, numerous pieces of evidence suggest that the risk of progression to active disease continues to decline substantially beyond the first 5-10 years after infection.^17^ For example, data on progression from longitudinal cohorts with *Mtb* infection in low-burden settings, along with models fit to cross-sectional data, are consistent with substantially declining progression rates over time.^18,19^ Furthermore, high rates of immunoreactivity reversion and low-to-moderate rates of progression among immunoreactive people after immunosuppression suggest that a meaningful proportion of *Mtb* infections clear over time. In people with cleared infections, immunoreactivity may indicate immunological memory rather than persistent infection with viable bacteria capable of causing reactivation disease.^20,21^

In this study, we sought to use mathematical models to understand the implications of our updated, more nuanced conceptualization of *Mtb* infection - in terms of estimating the importance of recent transmission in perpetuating TB epidemics in high-burden settings and projecting the impact of TB active case-finding.

## Methods

### Overview

We took a previously published dynamic compartmental *Mtb* transmission model (“Comparator Model”) and created an alternative version of the model structure, which differed only in the addition of a mechanism by which *Mtb* infections can clear (“Clearance Model”).^8^ We then calibrated both models to the same empirical epidemiological data from a high-burden setting (India). We subsequently used the calibrated models to project the impact of an illustrative active case-finding intervention, covering 70% of the population with a 65%-sensitive screening algorithm every 2 years. We compared simulated infection dynamics and projected intervention impact between the two models.

### Key Terms

We use the following definitions in this exercise:

- ***Mtb* Infection:** Exposure to *Mtb* at a previous point in life, resulting in a positive tuberculin skin test (TST) or interferon gamma release assay (IGRA) if tested, with ongoing risk of progressing to TB disease.
- **TB Disease:** Disease resulting in bacteriologically positive sputum, regardless of symptoms.
- **Immunoreactivity:** A positive TST or IGRA result, regardless of progression risk, prior TB disease, or infection status.
- **Cleared Infection:** Prior *Mtb* infection that is no longer at risk of progressing to TB disease. Cleared infections are assumed not to still generate immunoreactivity (an assumption we tested in sensitivity analysis).

### Model Structure

The Comparator Model followed a typical *Mtb* transmission model structure (Figure 1, Supplementary Methods, Supplementary Figure 1), in which a population transitions between six TB-related states. In this model, progression risk is highest immediately after *Mtb* infection (represented by a “recent infection” state with a higher progression rate) but persists for life among all who have been infected (represented by a “remote infection” state with a lower progression rate). In the alternative Clearance Model, we allowed for transition from the two infection states to a seventh, “cleared” state with no probability of progression (unless they experience reinfection). This clearance rate was calibrated.

**Figure 1.**
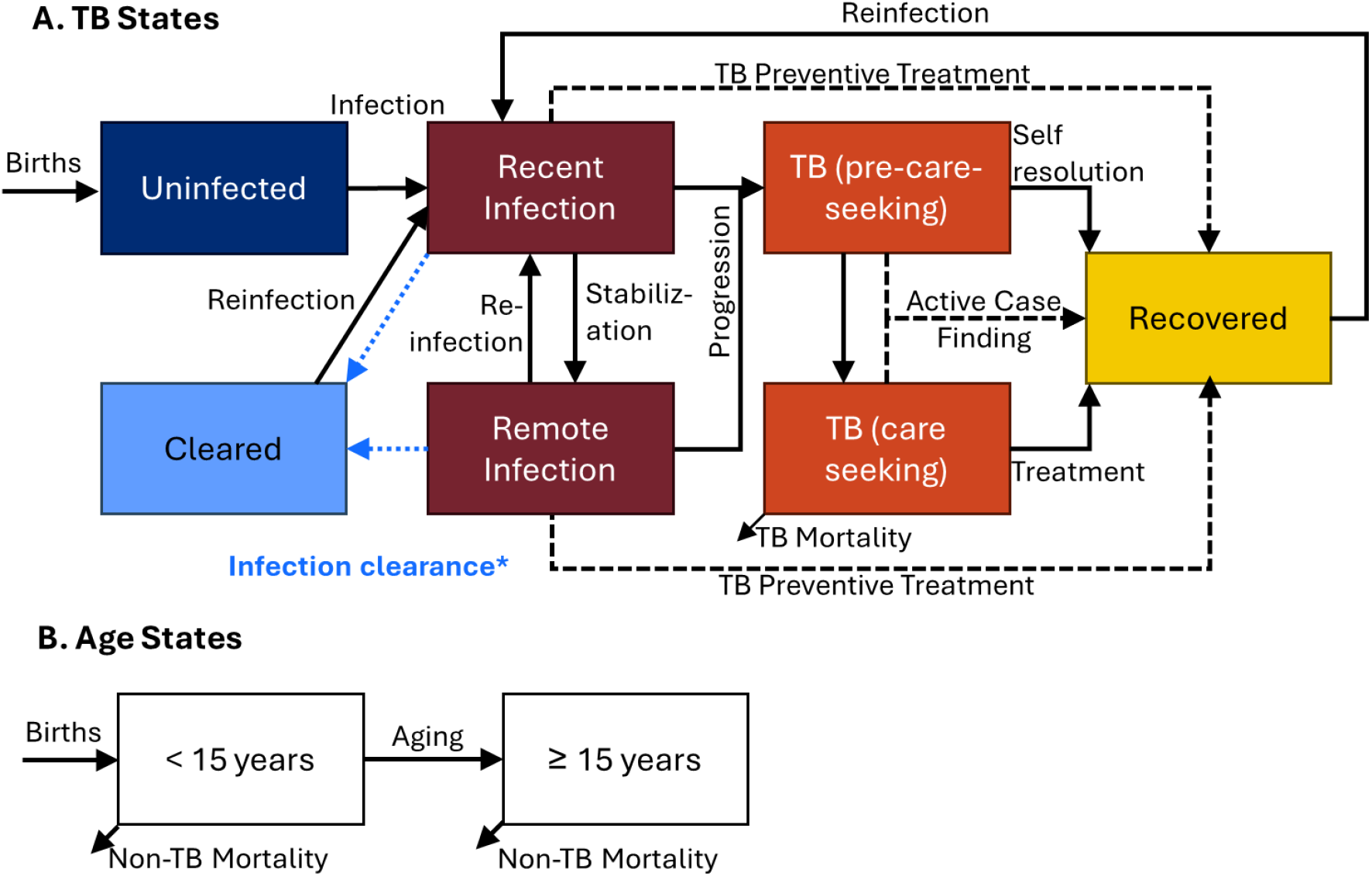
TB model structure. *Infection clearance (blue dashed arrows from Recent Infection and Remote Infection to Cleared) was included only in the Clearance model, not the Comparator model. In this model diagram, boxes represent TB-related (panel A) and age-related (panel B) states, while arrows represent transitions between states. Intervention-related transitions (resulting from active case-finding or provision of TB preventive treatment) are shown via black dashed lines. All individuals enter the model uninfected and in the younger (< 15 years) age stratum. TB mortality occurs from the care-seeking TB state only, while non-TB mortality occurs from all states. Apart from the birth and non-TB mortality rates and intervention-related parameters, all parameters were calibrated (details in the text). Additional details are available in the Supplementary Methods and Supplementary Figure 1. **Alt text:** Mathematical model diagram with two panels, in which boxes depict TB compartments (panel 1) and age compartments (panel 2) and arrows depict transitions between compartments.

In both models, we assumed that individuals with new TB disease start in a non-care-seeking phase, which can subsequently resolve to the recovered state or progress to symptom-driven care seeking. Individuals in the care-seeking phase can subsequently be diagnosed and treated/cured or die from TB; for simplicity in this model, we assumed that individuals who enter the care-seeking phase do not return to the non-care-seeking phase. Both models were further stratified into two age groups: children <15 years and adults ≥15 years, who face differential transmission, progression and mortality risks. Active case-finding and TPT interventions were simulated via movement to the recovered state from all TB disease states and all *Mtb* infection states, respectively.

### Model Calibration

Model parameters were calibrated to prevalence-survey-based and WHO-reported disease burden data from India (selected as an illustrative but representative high-burden country; Supplementary Methods, Supplementary Table 1), plus historical cohort data on progression over time after new *Mtb* infection (Table 1, Supplementary Tables 2-3). India-specific targets included WHO-estimates of TB incidence, the rate of change in incidence over time, and the case fatality ratio^22^, as well as estimates from India’s 2019-21 national TB prevalence survey of the prevalence of TB disease (in absolute terms and relative to annual notifications) and the prevalence of *Mtb* immunoreactivity.^23^

**Table 1.** Empirical data used as model calibration targets.

| Target | Empirical Estimate<br>[95% CI] | Comparator Model<br>Posterior<br>[95% UI] | Clearance Model<br>Posterior<br>[95% UI] | Sources for empirical estimates |
| --- | --- | --- | --- | --- |
| Annual incidence per 100,00 (2021) | 210 [178-244] | 193 [163-228] | 219 [187-251] | WHO Global TB Report <sup>22</sup> |
| Mortality to incidence ratio (2021) | 0.17 [0.14-0.21] | 0.17 [0.14-0.20] | 0.16 [0.13-0.20] |  |
| Annual % reduction in incidence (2002-21) | 2.9% [1.9-3.9%] | 0.6% [0.3-1.0%] | 2.6% [1.6-3.6%] |  |
| Prevalence per 100,000, ≥15 years (2019-21) | 316 [290-342] | 314 [285-341] | 312 [284-342] | India prevalence survey <sup>23*</sup> |
| Prevalence-to-notification ratio, ≥15 years (2019-21) | 2.84 [2.61-3.10] | 2.8 [2.6-3.1] | 2.8 [2.6-3.1] |  |
| Percent of prevalence pre-care-seeking, ≥ 15 years (2019-21) | 80% [73-88%] | 78% [69-85%] | 78% [71-84%] |  |
| Immunoreactivity prevalence, ≥ 15 years (2019-21) | 31% [20-44%] | 48% [40-57%] | 20% [14-29%] |  |
| Cumulative progression (first 2 years after infection) | 5.6% [4.3-7.1%] | 5.9% [4.6-7.2%] | 5.2% [4.0-6.5%] | Ferebee & Mount 1962; Sutherland 1968, reported in Menzies et al. 2018 <sup>24-26</sup> |
| Cumulative progression (years 3-10 since infection) | 1.5% [0.6-2.8%] | 2.8% [1.7-4.2%] | 1.8% [0.8-3.3%] |  |

\*The prevalence-to-notification ratio was calculated from a combination of prevalence survey data and national notification data.

Additional calibration targets were based on empirical estimates of time to progression to TB disease after infection, from two historical cohorts of TST-positive individuals with presumed new infection (based on recent exposure).^24–26^ We extracted estimates of cumulative progression in the first 2 years of infection and in years 3 through 10 after infection (chosen to inform recent and remote progression rates). Since these studies were conducted in low-prevalence settings, we assumed that all TB in these cohorts resulted from progression rather than reinfection.

After a burn-in period to reach a steady state, we induced declines in incidence over 20 years to match data on temporal trends in TB burden. In the main analysis, we used an annual linear increase in the treatment initiation rate to generate these declines, reflecting expansions in treatment that occurred over recent decades. In secondary analysis, we considered instead a declining transmission rate, reflecting development-related changes in opportunities for transmission or susceptibility to infection. The target for annual reductions in TB incidence was compared to the average rate of change over this 20-year period as estimated by WHO, and all other calibration targets were compared to model outputs at the end of these 20 years of incidence decline.

The models were coded in R v4.5.1 and calibrated using Bayesian Incremental Mixture Importance Sampling with uninformed priors (Supplementary Methods), resulting in 20,000 posterior parameter set samples per model.^27^ The model code is available at www.github.com/rycktessman/tb-model-clearance.

### Infection and Disease Dynamics

We first assessed key metrics of infection dynamics absent intervention, including the proportion of prevalent *Mtb* infections acquired recently, the proportion of TB incidence arising from recently versus remotely acquired infections, and the proportion of a given year’s TB incidence that could be attributed to transmission from the current population with prevalent TB disease. The latter was estimated by instantaneously moving all individuals with TB disease to the recovered compartment and running the model for one year. We also examined how key quantities not used in calibration, such as the annual risk of infection and the duration of TB disease, varied across the two models.

### Interventions

We used the calibrated models to simulate the impact of community-wide active case-finding campaigns conducted every 2 years, covering 70% of the population during each campaign with a 65% sensitive screening algorithm (Supplementary Methods; Supplementary Table 4). We evaluated intervention impact on TB incidence over a 10-year horizon and compared the projected impacts of case-finding between models. We also evaluated additional outcomes (prevalence, mortality) and considered a scenario in which TPT provision was added to the case-finding campaign.

### Sensitivity Analysis

We conducted probabilistic sensitivity analyses to evaluate uncertainty across all model parameters. Main results are presented as mean values, with 95% uncertainty intervals (UI) representing the 2.5^th^ and 97.5^th^ percentiles across 20,000 parameter set samples. We also evaluated correlations between parameters and modeled outcomes, examined variation in projected intervention impact between the lowest and highest deciles of each parameter’s distribution (as a form of multivariable one-way sensitivity analysis), and conducted two-way sensitivity analysis on the percentages of TB disease and *Mtb* infection cured via intervention.

## Results

### Model Calibration

Both models achieved reasonable fits to most of the nine empirical calibration targets (Table 1, Supplementary Figure 2). The Comparator Model was unable to replicate pre-intervention declines in incidence and tended to overestimate immunoreactivity prevalence and late progression, while the Clearance Model (when assuming no immunoreactivity among people with cleared infections) estimated somewhat lower immunoreactivity prevalence than measured by the prevalence survey.

The calibrated effective contact rate (“beta”) was 4 times higher in the Clearance Model than the Comparator Model, suggesting more transmission is required to achieve consistency with epidemiological targets when some individuals clear their infections (Supplementary Table 5). In the Clearance Model, the clearance rate corresponded to approximately half of people who had not yet progressed clearing their infection within 5 years. In both models, the recent infection period lasted an average of 5-6 months, but the proportion of progression that occurred during that period was greater in the Clearance Model.

### Infection and Disease Dynamics

Recent infection accounted for a greater share of total *Mtb* infection prevalence under the Clearance Model (8% [4-15%]) versus the Comparator Model (1% [1-2%]; Figure 2). Since progression rates from recent infection are higher than remote reactivation rates, this resulted in a substantially greater percentage of TB incidence attributable to progression from recent infection in the Clearance Model: 61% [35-79%], versus 28% [14-46%] in the Comparator Model. Similarly, under the Clearance Model, 32% [20-44%] of incidence in a given year resulted from transmission from the current population with prevalent TB (i.e., at the beginning of that year), versus 14% [8-23%] under the Comparator Model.

**Figure 2.**
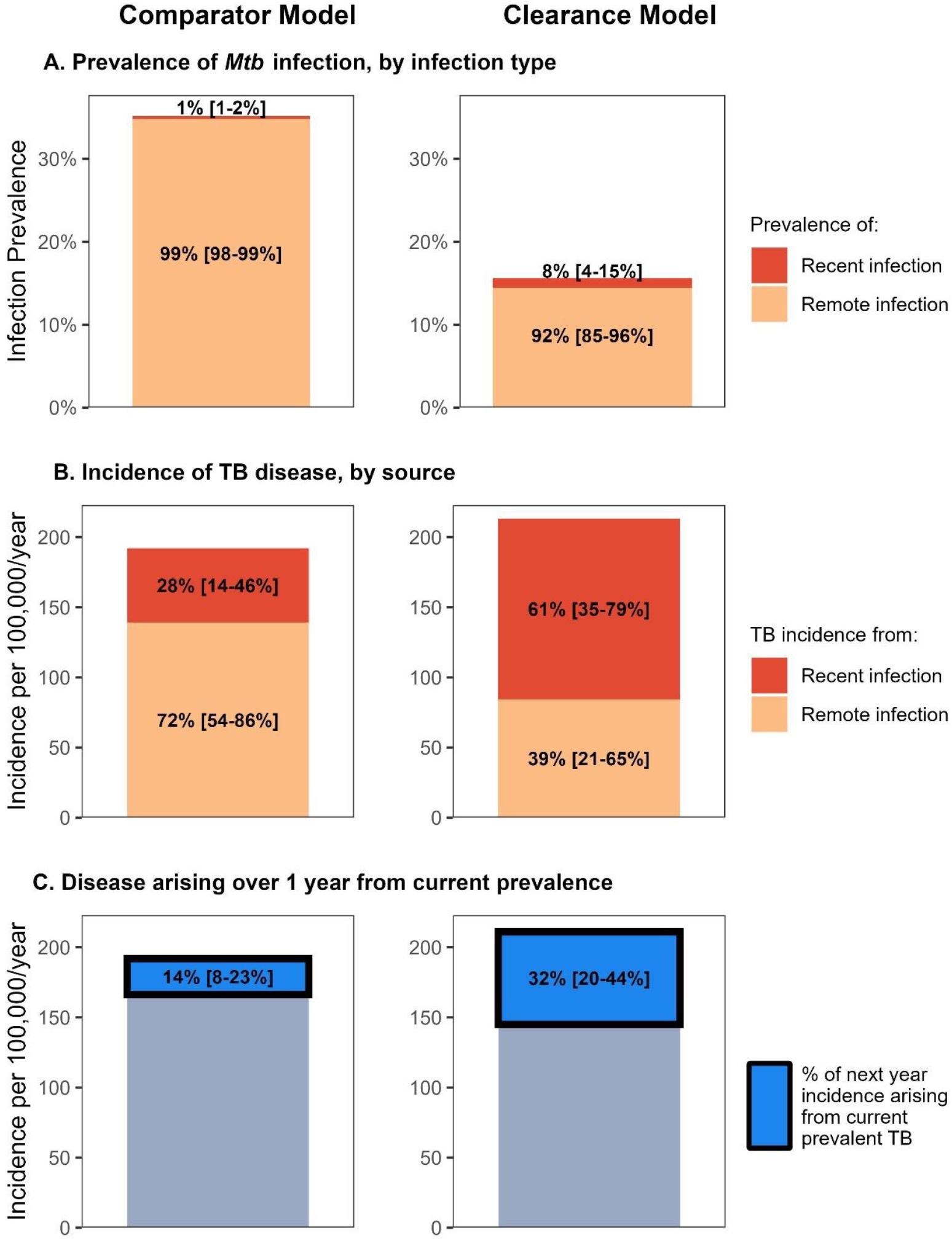
Effect of considering immune clearance on modeled TB infection dynamics. This figure illustrates how inclusion of *Mtb* infection clearance affects modeled infection and disease dynamics. Panel A demonstrates that, without a mechanism for immune clearance (“Comparator Model”), people who were infected remotely account for a larger absolute population, and also a larger fraction of those with *Mtb* infection. Because calibration data shared by both models results in similar progression rates after infection, a greater prevalence of recent *Mtb* infection results in the Clearance model estimating that a larger proportion of incidence arises from recent (vs. remote) infection than the Comparator model (Panel B). Since interventions to diagnose and treat people with current TB disease will primarily affect the burden of recent infection, the Clearance model estimates a larger impact from such interventions (Panel C). In panel C, the “disease arising over 1 year from current prevalence” represents the fraction of new incident disease in a given year that would be averted if all people with TB disease at the start of that year were instantaneously diagnosed and treated at year-start. All results show means across 20,000 model simulations, with corresponding 95% uncertainty intervals in parentheses. **Alt Text:** Bar chart with 3 panels (organized horizontally) and 2 columns (first column shows results under the Comparator model without clearance; second column shows results under the Clearance model). The top panel displays the prevalence of *Mtb* infection, with shading indicating the proportion of that prevalence that is recent vs. remote infection. The middle panel displays TB incidence, with shading indicating the proportion of that incidence that is from progression of recent infection vs. reactivation of remote infection. The bottom panel again displays TB incidence, with the proportion of incidence that could be averted over the next year by finding and treating all people with current prevalent TB emphasized via bright blue shading and a thick black outline. Text labels show numerical estimates on each panel (means, with 95% uncertainty intervals in brackets).

Consistent with infection from recent transmission playing a greater role when clearance is considered, the annual risk of infection was also higher under the Clearance Model (4.8% [2.5-7.3%]) than the Comparator Model (1.1 % [0.8-1.5%]) (Supplementary Table 6). The duration of TB disease, estimated both with current treatment intensity and absent any treatment, was similar under both models (Supplementary Table 7).

### Intervention Impact

Screening 70% of the population every 2 years with a 65% sensitive screening algorithm was projected to yield a significantly greater reduction in incidence by year 10 under the Clearance Model (45% [28-57%]) than the Conventional Model (11% [6-18%]) (Figure 3). For the combined intervention of both active case-finding and TPT, impact was similar across models (all 95% UIs within 79-85%; Supplementary Figure 3). Corresponding prevalence and mortality estimates over time are shown in Supplementary Figures 4-5.

**Figure 3.**
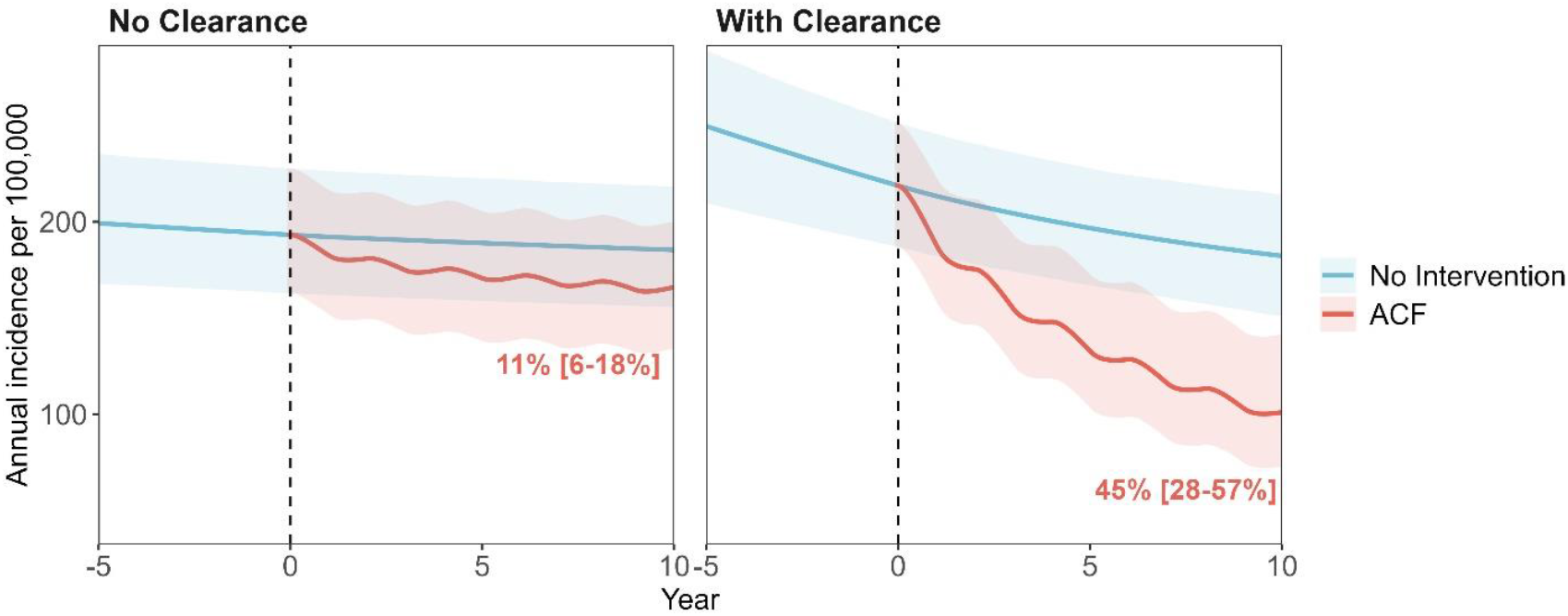
Projected impact of active case-finding under models with and without infection clearance. Figure shows the projected the mean annual TB incidence per 100,000 people (solid lines), with 2.5th and 97.5th percentiles (shaded ribbons), over ten years of simulated interventions (years 0 through 10), including no intervention (blue) and Active Case-Finding/ACF waves every 2 years (red). Despite overlapping uncertainty ribbons due to variation in baseline incidence across posterior parameter sets, all simulations projected a reduction in incidence from ACF (vs. no intervention). Text labels indicate the percent reduction in incidence at year 10, relative to no intervention (text shows means, with 95% uncertainty intervals in brackets). Panels differ by whether the model structure includes infection clearance (panel A = Comparator Model without clearance; panel B = Clearance Model). Incidence trends in the 5 years preceding the intervention period are also shown in each panel (years −5 through 0). **Alt text:** Line graphs in 2 panels; the left panel shows results under the Comparator Model without clearance and the right panel shows results under the Clearance Model. Each panel shows incidence (vertical axis) over time (horizontal axis) with no additional interventions (blue line) or with active case-finding campaigns every 2 years (red line). Text labels on each panel indicate the reduction in incidence after 10 years from the active case-finding campaigns compared to no intervention (means with 95% uncertainty intervals in brackets).

### Sensitivity Analyses

We conducted sensitivity analyses that varied the mechanism of pre-intervention incidence declines (from treatment-to susceptibility-mediated) and the proportion of cleared infections that remain immunoreactive (from 0 to 100%; calibration data shown in Supplementary Figures 6-7). Across these analyses, infection from recent transmission continued to make up a greater share of *Mtb* infection prevalence and TB disease incidence under the Clearance Model than the Comparator Model, and annual risk of infection was consistently higher (Supplementary Tables 6-7). The greater projected impact from active case-finding under the Clearance Model was also consistent across these sensitivity analyses (Supplementary Fig 8-9), and was similarly consistent when varying the coverage and efficacy of case-finding (Supplementary Figure 10). The impact of case-finding on incidence was most sensitive to the progression rate from remote infection; other influential parameters included the effective contact rate, the spontaneous resolution rate, and the treatment rate (Supplementary Figures 11-12).

## Discussion

A more nuanced conceptualization of *Mtb* infection that allows for the possibility of infection clearance is starting to be incorporated into mathematical models,^12^ and studies have explored its implications for natural history trajectories^16^, lifetime risk of disease,^28^ and vaccine trials^14^. Here, we expand on this understanding by demonstrating the implications for disease dynamics and the impact of interventions. We found that incorporating clearance into a typical *Mtb* transmission model is consistent with a greater role of recent transmission in sustaining TB epidemics, implying that interventions which interrupt that transmission – for example, by finding more people with TB and linking them to effective treatment – could have more substantial and lasting epidemiological impact than previously estimated. Importantly, mass TPT provision enhanced impact in both models, but was not necessary to achieve large reductions in incidence under the Clearance Model.

To understand these results, one must consider how models that do not include infection clearance must operate to fit observed data. Since a large fraction of the population is assumed to have ongoing, lifelong reactivation risk, the majority of incident TB must derive from the pool of remote infections. This pool cannot be reduced by transmission-reducing interventions like case-finding. By contrast, if a substantial fraction of infections clear over time, a greater fraction of TB incidence arises from recent transmission - which can be interrupted by interventions like case-finding. This behavior also explains why the Clearance Model was better able to replicate empirical estimates of declines in incidence that have occurred over the last 20 years than the Comparator Model, under which it is difficult to achieve incidence reductions absent interventions that reduce progression. Importantly, while we chose to model infection clearance, any mechanism that continues to reduce the risk of progression to disease beyond the first few years of infection, or shifts the balance of progression from remote to recent infection in other ways, is expected to have a similar effect.

This exercise was not intended to provide precise estimates of the impact of any specific case-finding intervention. Findings regarding the greater importance of recent transmission and greater impact of case-finding when clearance is considered were consistent across a range of intervention coverages and efficacies, demonstrating that our conclusions are not sensitive to intervention-related parameters. As anticipated, we projected a greater impact from case-finding than previous models that did not include a clearance (or similar) mechanism.^8–11^ The annual risk of infection estimated by the Clearance Model exceeded an estimate from India based on TST surveys among children.^29^ However, a recent review found that cross-sectional surveys among children likely underestimate the true annual risk of infection in high-burden settings, which may be closer to 5-10% (consistent with our estimates from the Clearance Model).^30^

Although we calibrated to data from India, these findings are expected to generalize to other high-burden settings with low HIV prevalence. The balance of key targets that influenced model behavior - immunoreactivity (or infection) prevalence, incidence, and progression by time since infection – are similar in other high-TB-burden countries (Supplementary Table 1). Some favor recent infection even more than India’s data – with higher incidence, a lower ratio of infection prevalence to incidence, and/or greater annual reductions in incidence (where adequate data exist to inform trend estimates).^22,31^ Other unique epidemiologic features of India’s TB epidemic, such as a high prevalence-to-notification ratio, were not major drivers of calibration fit or projected intervention impact. On the other hand, these results may not generalize to settings with high TB-HIV co-prevalence. A setting-specific model that represents TB-HIV dynamics would be needed to apply the structural questions considered here to a high HIV-prevalence setting.

Our findings should be interpreted in light of certain limitations. First, our main analysis assumed that cleared infections no longer exhibit immunoreactivity. While the percentage of cleared infections that maintain immunoreactivity remains uncertain, we found consistently higher (>3x) impact of case-finding under the Clearance Model regardless of this percentage (varied from 0% to 100% in sensitivity analysis). Second, our model’s TB disease states only captured individuals with sputum-positive TB. Alternative model structures that explicitly represent a sputum-negative TB disease state exhibit different underlying dynamics (such as a higher clearance rate than estimated here)^16^ but similarly project a greater impact of active case-finding than many other modeling exercises.^12^ Finally, all models are inherently simplified representations of complex dynamics - but we believe such simplified representations are useful when trying to isolate the effect of a single mechanism, such as that of infection clearance explored here.

Numerous pieces of evidence suggest that, rather than *Mtb* infection representing a protracted state of persistent reactivation risk, many people either clear their *Mtb* infections or stabilize to a very low risk of reactivation within a relatively short time after infection. This fundamental reconceptualization of *Mtb* infection dynamics has important implications for the impact of transmission-reducing interventions. Specifically, remote infections likely play less of a role in driving the ongoing burden of TB in high-incidence settings than has previously been thought, implying that interventions that reduce transmission (e.g., case-finding) may have greater impact than previously estimated. These findings support the continued rollout of setting-specific community case-finding efforts, along with targeted programs delivered to people at high risk of progression (such as household contacts, people with HIV, and people with undernutrition), as part of a comprehensive intervention package to reduce the global burden of TB.

## Acknowledgements

We would like to thank Nicolas Menzies for sharing a copy of the Sutherland et al. study (reference 26), data from which was used to quantify the early and late progression targets to which we calibrated the model.

## Author Contributions

TSR, EAK, and DWD conceptualized the study, and SS, ATF, and PK contributed to initial discussions on motivation and scope. TSR curated and cleaned the data, ran all analyses, and wrote the first draft of the manuscript. SS provided initial model code that was adapted by TSR. EAK provided ongoing input and suggestions on methods and preliminary analyses. EAK, DWD, SS, ATF, and PK contributed to discussions of preliminary results and subsequent model drafts. EAK and TSR verified the study data. All authors had full access to all the data in the study and accept responsibility to submit for publication.

## Data Availability

Model code is published online at www.github.com/rycktessman/tb-model-clearance. All data used in the study are displayed and cited in Table 1, Supplementary Table 3, and Supplementary Table 4.

## Meetings

Findings from this study were presented at the Union World Conference on Lung Health, Copenhagen, November 20, 2025.

