## Supplemental Material for "Modeling the impact of case finding for tuberculosis: The role of infection dynamics"

Supplementary Figure 3: Projected impact of case finding and mass TPT provision on incidence ... 14

Supplementary Figure 4: Projected impact of case finding and mass TPT provision on prevalence . 15

Supplementary Figure 5: Projected impact of case finding and mass TPT provision on TB mortality 16

**Supplementary Methods**

*Model Details*

Our model, which was adapted from a previously case finding and TPT modeling exercise (1), follows a typical *Mtb* transmission model structure but includes a mechanism for infection clearance (Fig. S1). Our simulated population transitions between 7 TB-related states: one uninfected state ($U$), *two Mtb* infection states (recent infection $I_1$ and remote infection $I_2$), two TB disease states (a pre-care-seeking state $A_1$ and a care seeking state $A_2$), a recovered state ($R$), and a cleared state ($C$). The model is further stratified into two age groups (defined by the subscript $a$): children under fifteen years ($a = 1$) and adults aged fifteen years and older ($a = 2$), who face differential progression and mortality risks.

The population is born uninfected and fully susceptible to infection. Infections occur at rate $\lambda U_a$, where the force of infection $\lambda$ is a function of the current adult disease prevalence (children are assumed to not be infectious) and effective contact rate, $\beta$ (see equations below).

Once infected, progression to disease is higher among those recently infected ($p > q$) and varies by age (via the relative risk parameter $x_a$, where $x_1$ is calibrated and $x_2 = 1$). Stabilization from recent to remote infection is also calibrated (the average duration of time spent in the recent infection state is therefore effectively $1/s$). In the Comparator Model (without clearance), those with *Mtb* infection face a lifelong risk of progression to active TB (i.e., disease). In the Clearance Model, those with remote infection can clear at an annual rate of $c$, at which point they transition to the cleared state, where in the main analysis they are assumed to no longer test positive on a TB immunoreactivity test (e.g., TST or IGRA; this assumption was varied in sensitivity analysis). Reinfection (transition from the remote infection or recovered states to the recent infection state) can occur, but those with prior infection are assumed to retain some protection from reinfection (via the $\chi$ parameter, which represents the relative risk of reinfection after initial infection).

Those who have progressed to TB disease start in a non-care-seeking phase, during which they can spontaneously resolve ($w$) to the recovered state or progress to develop symptoms significant enough to seek care ($r$). Care seeking eventually results in either treatment and cure, or death (TB-specific mortality rate = $\mu_{TB}$). The rate of cure is a product of the detection and treatment initiation rate, $\omega$, and the treatment success ratio, $k$.

Death from non-TB-related causes occurs from all states. Birth ($b$) and background mortality rates ($\mu_a$) are based on demographic data from India (2) and do not vary over time. Children transition to the respective adult compartments at a rate of $1/15$. All other parameters are calibrated.

In this model, we simulated active case-finding and TB preventive treatment (TPT) via movement to the recovered state from the TB disease and *Mtb* infection states, respectively (indicated via dashed black lines in Fig. S1). The active case-finding rate, $ACF$, is a product of screening coverage, screening and confirmatory test sensitivity, and treatment success. The TPT rate is a product of screening coverage, TST coverage (e.g., the proportion of people with *Mtb* infection who have their TST read and test positive), TPT acceptance and completion, and TPT efficacy.

*Model Equations*

In the below equations, curly brackets { } designate an indicator function, which is equal to 1 if the statement inside the brackets is true, and 0 otherwise. The subscript $a$ refers to age group. Clearance (clearance rate = $c$) is only modeled in the Clearance Model; it is fixed at 0 in the Comparator Model. See Fig. S1 for additional information.

$$\frac{dU_a}{dt} = b\{a = 1\} + \frac{1}{15}U_1\{a = 2\} - \left(\lambda + \frac{1}{15}\{a = 1\} + \mu_a\right)U_a$$

$$\frac{dI_{1,a}}{dt} = \lambda U_a + \chi\lambda(I_{2,a} + R_a + C_a) + \frac{1}{15}I_{1,1}\{a = 2\} - \left(s + x_a p + c + TPT_a + \frac{1}{15}\{a = 1\} + \mu_a\right) I_{1,a}$$

$$\frac{dI_{2,a}}{dt} = sI_{1,a} + \frac{1}{15}I_{2,1}\{a = 2\} - \left(\chi\lambda + x_a q + c + TPT_a + \frac{1}{15}\{a = 1) + \mu_a\right) I_{2,a}$$

$$\frac{dA_{1,a}}{dt} = x_a pI_{1,a} + x_a qI_{2,a} + \frac{1}{15}A_{1,1}\{a = 2\} - \left(w + r + ACF_a + \frac{1}{15}\{a = 1\} + \mu_a\right) A_{1,a}$$

$$\frac{dA_{2,a}}{dt} = rA_{1,a} + \frac{1}{15}A_{2,1}\{a = 2\} - \left(k\omega + ACF_a + \frac{1}{15}\{a = 1\} + \mu_{TB} + \mu_a\right) A_{2,a}$$

$$\frac{dR_a}{dt} = wA_{1,a} + k\omega A_{2,a} + ACF_a\left(A_{1,a} + A_{2,a}\right) + TPT_a\left(I_{1,a} + I_{2,a}\right) + \frac{1}{15}R_1\{a = 2\} - \left(\chi\lambda + \frac{1}{15}\{a = 1\} + \mu_a\right) R_a$$

$$\frac{dC_a}{dt} = c\left(I_{1,a} + I_{2,a}\right) + \frac{1}{15}C_1\{a = 2\} - \left(\chi\lambda + \frac{1}{15}\{a = 1\} + \mu_a\right) C_a$$

Where:

$$\lambda = \beta(A_{1,2} + A_{2,2})$$

$$ACF = screening\_coverage * test\_sensitivity * k$$

$$TPT = screening\_coverage * TST\_coverage * TPT\_acceptance * TPT\_completion * TPT\_efficacy$$

Declines in incidence over time in the pre-intervention period were either induced via annual linear increases in the treatment initiation rate (main analysis) or declines in the effective contact rate (sensitivity analysis). In those respective variations, these parameters were modified as follows, for a given year $t$:

$$\omega_t = \omega * \left(1 + change_{omega}\right)^t$$

$$\beta_t = \beta * \left(1 - change_{\beta}\right)^t$$

Reductions in the effective contact rate continued beyond the burn-in period (into the intervention period), while treatment initiation rates were assumed to plateau at their maximum at the end of the burn-in period (further treatment rate increases did not continue into the intervention period)

*Country Selection*

India was selected as an illustrative country to model in this exercise because it is an important country epidemiologically (largest number of people with TB in the world), it has a high TB incidence (annual incidence 210 per 100,000), and it has a relatively low burden of HIV (prevalence 0.2%) – since modeling a setting with high HIV prevalence which would require representing HIV in a way that would add complexity to our model structure (3). India is also fairly representative in terms of several empirical estimates that were used in the calibration process (Table S1).

*Calibration Details*

The model was calibrated to empirical targets (Table S2) using Bayesian Incremental Mixture Importance Sampling (via an adaption to the R package *IMIS*) (4, 5). Specifically, for each model we randomly sampled 20 sets of 2,000 parameter sets each from relatively uninformed prior distributions (Table S3), and then conducted 100 rounds with targeted sampling of 100 parameters each based on the highest-likelihood parameter sets from initial sampling, saving 20 posterior sets of 1000 posterior parameter sets (by sampling with replacement using posterior probability-based sampling weights) for a combined posterior distribution of 20,000 parameter set samples.

*Intervention Details*

The modeled interventions were simulated as waves of population-level active case-finding every 2 years (starting in the first year of the post-burn-in intervention period), with or without provision of TPT for people with *Mtb* infection (among whom TB disease was assumed to have been ruled out). For simplicity, intervention coverage, diagnostic sensitivity, linkage to care, and intervention efficacy were assumed not to vary by age. Modeled outcomes, such as incidence, prevalence, and mortality were compared against a "No Intervention" scenario with no active case-finding and no TPT.

Intervention-related parameters were based on the published literature and a previous active case-finding and TPT modeling exercise (1), and are shown in Table S4. Parameters related to intervention coverage were selected to be ambitious but feasible in a high-burden context, and are based on data recently reported from a mass screening program in the Marshall Islands by Ragonnet and colleagues (6).

In order to focus the analysis on differences related to model structure, we did not consider uncertainty in the intervention parameters in the main analysis. Instead, we conducted two-way sensitivity analysis on the proportion of TB disease and *Mtb* infection cured under each intervention wave.

**Supplementary Figure 1: Detailed model diagram**

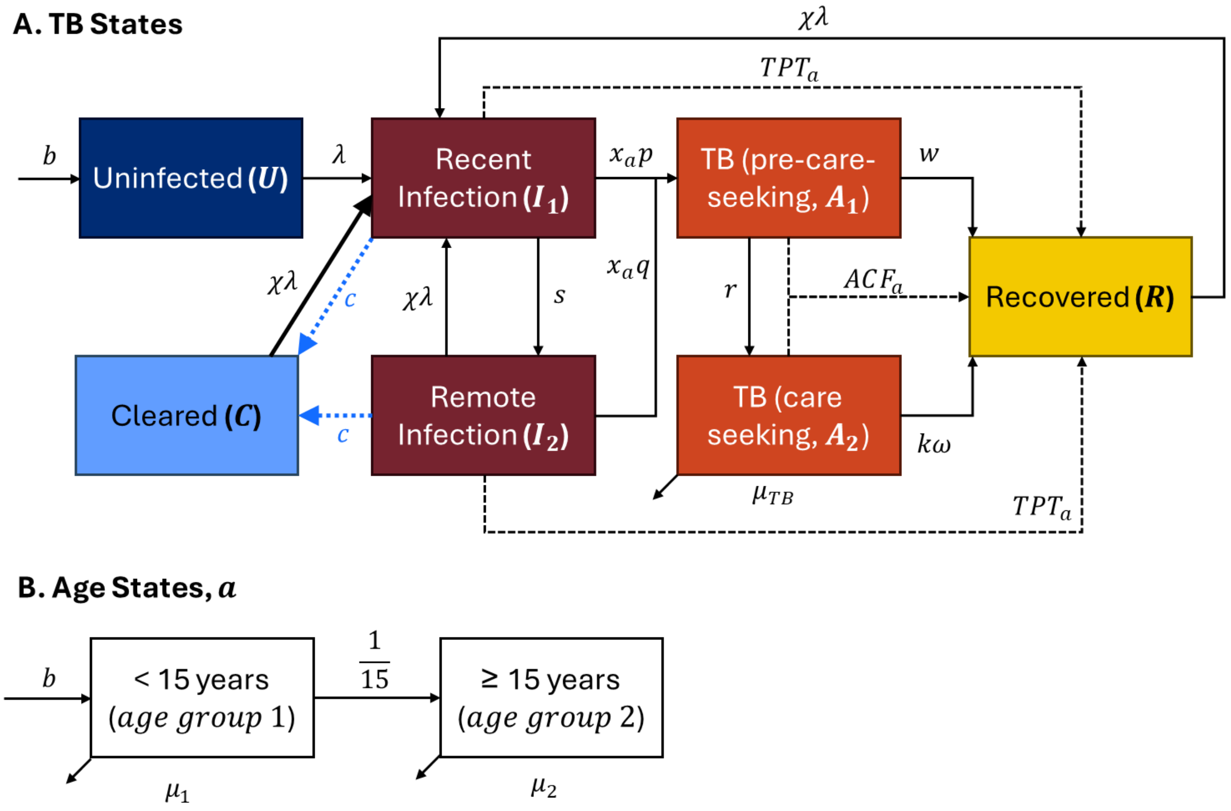

*Infection clearance (blue dashed arrows from Recent Infection and Remote Infection to Cleared) was included only in the Clearance model, not the Comparator model.

In this model diagram, boxes represent TB-related (panel A) and age-related (panel B) states, while arrows represent transitions between states. Intervention-related transitions (resulting from active case-finding or provision of TB preventive treatment) are shown via black dashed lines. All individuals enter the model uninfected and in the younger (< 15 years) age stratum. TB mortality occurs from the care-seeking TB state only, while non-TB mortality occurs from all states. Apart from the birth and non-TB mortality rates and intervention-related parameters, all parameters were calibrated (details in the text). See Supporting Text 1 ("Model Details") for additional information.

**Supplementary Table 1: Empirical burden data from 30 high-TB-burden countries**

| Country | Annual incidence per 100,000 (2021)* | *Mtb* infection prevalence (2016)† | Ratio (# people with *Mtb* infection per annual # people with incident TB) | Annual change in incidence* |
|---|---|---|---|---|
| Angola | 325 | 23% | 69 | 0.1% |
| Bangladesh | 221 | 29% | 129 | 0.0%** |
| Brazil | 48 | 13% | 276 | -0.4% |
| Central African Republic | 540 | 47% | 88 | 0.0%** |
| China | 55 | 26% | 474 | -2.9% |
| Congo | 370 | 20% | 55 | -0.4% |
| DPR Korea | 513 | 36% | 71 | 0.0%** |
| DRC Congo | 318 | 28% | 89 | -0.1% |
| Ethiopia | 119 | 24% | 200 | -5.8% |
| Gabon | 513 | 29% | 56 | -0.6% |
| India | 210 | 27% | 130 | -2.4% |
| Indonesia | 354 | 48% | 135 | -0.2% |
| Kenya | 251 | 13% | 50 | -3.7% |
| Lesotho | 614 | 35% | 57 | -2.5% |
| Liberia | 308 | 23% | 76 | 1.0% |
| Mongolia | 428 | 40% | 93 | 0.0%** |
| Mozambique | 361 | 35% | 98 | 0.3% |
| Myanmar | 360 | 47% | 132 | -2.1% |
| Namibia | 457 | 38% | 83 | -4.7% |
| Nigeria | 219 | 18% | 84 | 0.0%** |
| Pakistan | 264 | 28% | 106 | -0.2% |
| Philippines | 424 | 35% | 83 | -0.1% |
| Papua New Guinea | 650 | 40% | 61 | 0.7% |
| Sierra Leone | 289 | 28% | 98 | -0.4% |
| South Africa | 513 | 32% | 61 | -3.1% |
| Thailand | 143 | 34% | 236 | -2.6% |
| Uganda | 199 | 15% | 74 | -1.3% |
| Tanzania | 208 | 30% | 142 | -4.4% |
| Viet Nam | 173 | 39% | 226 | -2.4% |
| Zambia | 307 | 32% | 103 | -4.0% |

Table shows key burden indicators for all countries classified as high-TB-burden by the WHO in 2021-25.

*Estimates from the WHO Global TB report (2023) (3);

† Estimates from Houben and Dodd (2016) (7). Note that we used an alternate (higher) estimate of immunoreactivity prevalence for India from the 2019-21 prevalence survey, which was 31% [20-44%]. Recent survey-based estimates were not available for most countries, and so for comparability across countries we present the estimates from Houben and Dodd instead.

**Settings with a 0.0% trend in incidence generally lacked data to inform estimates of changing incidence over time.

**Supplementary Table 2: Calibration targets and distributions**

| Target | Value [95% CI] | Distribution used to calculate likelihood | Sources |
|---|---|---|---|
| Annual incidence per 100,00 | 210 [178-244] | Normal (210.0, 16.8) | WHO Global TB Report (3) |
| Mortality to incidence ratio | 0.17 [0.14-0.21] | Beta (75.1, 366.4) | |
| Annual % reduction in incidence* | 2.9% [1.9-3.9%] | Normal (0.029, 0.005) | |
| Prevalence per 100,000, ≥ 15 years | 316 [290-342] | Normal (316.0, 13.3) | India prevalence survey (8) |
| Prevalence to notification ratio, ≥ 15 years | 2.84 [2.61-3.10] | Normal (2.8, 0.13) | |
| % prevalence pre-care-seeking, ≥ 15 years | 80% [73-88%] | Beta (67.3, 16.8) | |
| Immunoreactivity prevalence, ≥ 15 years | 31% [20-44%] | Beta (16.7, 36.5) | |
| Cumulative progression (first 2 years after infection) | 5.6% [4.3-7.1%] | Beta (58.7, 982.9) | Ferebee & Mount 1962 and Sutherland 1968, reported in Menzies et al. 2018 (9–11) |
| Cumulative progression (years 3-10 since infection) | 1.5% [0.6-2.8%] | Beta (7.5, 484.7) | |

Distributions (column 3) are shown as (mean, standard deviation) for normal distributions and (alpha, beta) for beta distributions, where the mean of the beta distribution equals alpha/(alpha + beta). Distributions were generally selected to fit the means and confidence intervals shown in column 2.

**Supplementary Table 3: Prior parameter distributions**

| Parameter | Abbreviation in equations | Prior range | Prior range source |
|---|---|---|---|
| TB mortality rate | $\mu_{TB}$ | 0.05, 0.75 | Uninformed |
| Effective contact rate | $\beta$ | 3, 30 | Uninformed |
| Relative risk of reinfection | $\chi$ | 0, 1 | Uninformed |
| Relative rate of progression, < 15s | $x_1$ | 1.0, 1.6 | Martinez et al. 2020 (12) |
| Early progression rate, adults | $p$ | 0.01, 0.2 | Uninformed |
| Late progression rate, adults | $q$ | 0.00001, 0.02 | Uninformed |
| Clearance rate* | $c$ | 0, 0.25 | Uninformed |
| Transition rate to care-seeking | $r$ | 0.1, 1 | Uninformed |
| Spontaneous resolution rate | $w$ | 0.1, 1 | Uninformed |
| Treatment initiation rate | $\omega$ | 0.1, 1.5 | Uninformed |
| Treatment success ratio | $k$ | 0.6, 0.97 | Uninformed |
| Annual increase in treatment initiation rate** | $change_{\omega}$ | 0.01, 0.3 | Uninformed |
| Annual decline in effective contact rate** | $change_{\beta}$ | 0.0005, 0.05 | Uninformed |

All parameters were sampled from uniform distributions, with the minima and maxima of each distribution shown in the table. Some uninformed priors were narrowed after initial calibration in order to increase efficiency.

*The clearance rate was only calibrated in the Clearance Model (it was fixed at 0 in the Comparator Model).

**In the main analysis, annual decline in effective contact rate was set to 0. In sensitivity analysis, this was calibrated and instead the annual increase in the treatment initiation rate was set to 0. These 2 parameters allowed for 2 different methods to induce declines in incidence during the pre-intervention period.

**Supplementary Table 4: Intervention parameters**

| Parameter | Value | Source and Notes |
|---|---|---|
| Screening coverage | 70% | Estimate of reasonable coverage from Shrestha et al. 2021 (1); conservatively lower than the 81-85% reported in Ragonnet et al. 2022 to account for multi-year screening (6) |
| Screening + test sensitivity | 65% | Based on Zifodya et al. 2021 and Shapiro et al. 2021 (13, 14) |
| TST sensitivity | 90% | Jonas et al. 2023 (15). Defined as sensitivity relative to IGRA, since immunoreactivity prevalence in the model is based on IGRA positivity. |
| % people *with Mtb* infection who have their TST read and test positive | 86.5% | Ragonnet et al. 2022 (6) |
| TPT acceptance/initiation | 85% | Ragonnet et al. 2022 (6) |
| TPT completion | 85% | Ragonnet et al. 2022 (6) |
| TPT efficacy | 69% | Pease et al. 2017 (16) |

**Supplementary Figure 2: Fit of Conventional and Clearance models to 9 calibration targets (main analysis)**

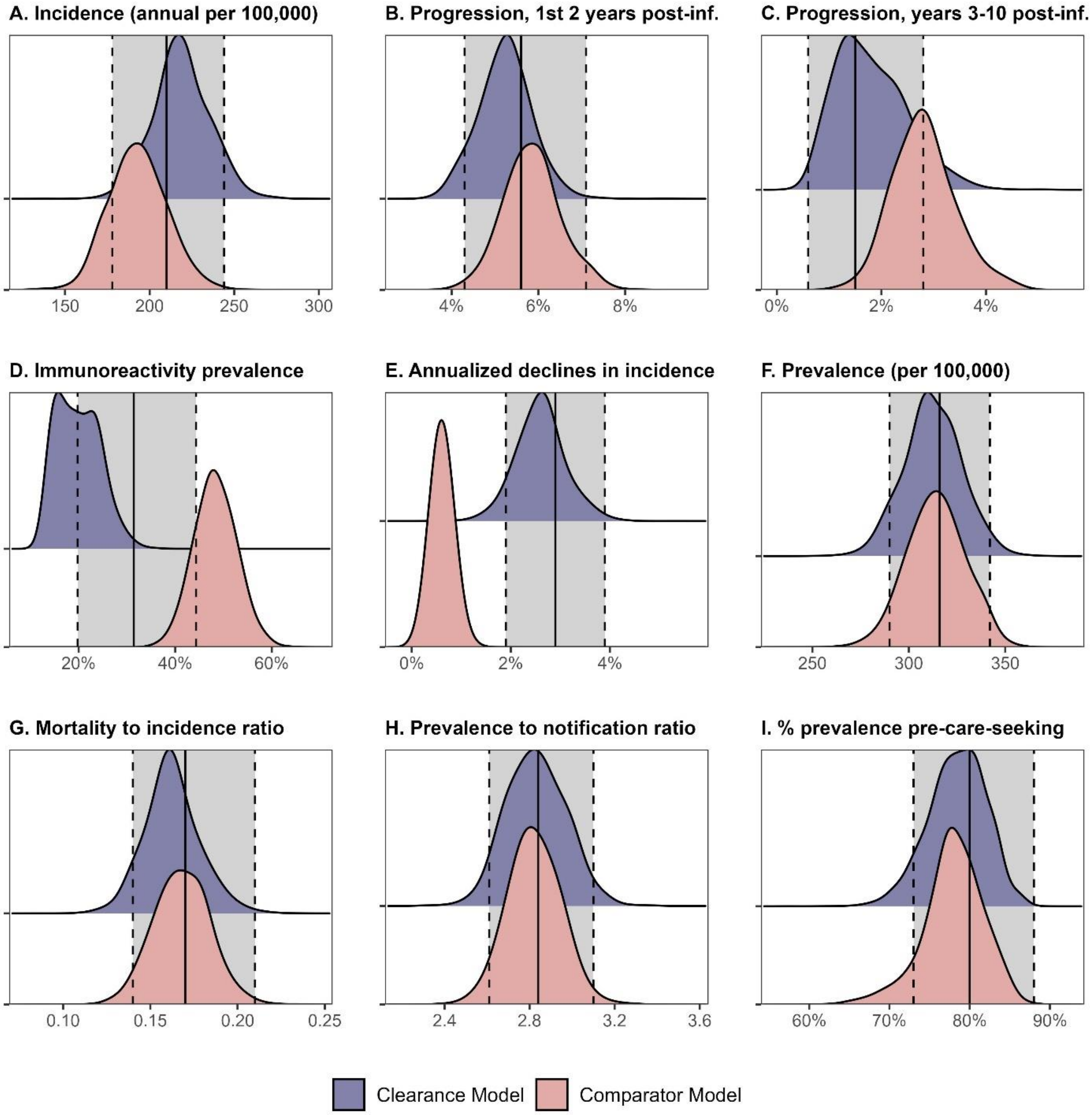

Figure shows the posterior distributions of 9 modeled outputs (shaded density plots) against corresponding calibration targets (mean values indicated by solid black vertical lines, 95% confidence intervals indicated by grey shading with dashed vertical lines. Outputs from the Clearance Model are shown in blue and outputs from the Comparator Model (no clearance) are shown in red.

**Supplementary Table 5: Posterior parameter distributions**

| Parameter | Abbreviation in equations | Comparator Model | Clearance Model |
|---|---|---|---|
| Effective contact rate | $\beta$ | 4.6 [3.4-6.1] | 19.4 [9.8-29.1] |
| Early progression rate | $p$ | 0.13 [0.07-0.19] | 0.11 [0.06- 0.17] |
| Late progression rate | $q$ | 0.004 [0.002-0.006] | 0.006 [0.002- 0.016] |
| Stabilization rate | $s$ | 2.3 [1.1-3.4] | 2.2 [1.2- 3.2] |
| Clearance rate | $c$ | - | 0.15 [0.08- 0.24] |
| Spontaneous resolution rate | $w$ | 0.40 [0.25-0.63] | 0.44 [0.32- 0.59] |
| Transition to care seeking rate | $r$ | 0.46 [0.38-0.56] | 0.39 [0.31- 0.47] |
| TB mortality rate | $\mu_{TB}$ | 0.52 [0.35-0.71] | 0.49 [0.32- 0.67] |
| Treatment rate | $\omega$ | 0.42 [0.18-0.81] | 0.34 [0.17- 0.61] |
| Relative rate of progression, < 15s | $x_1$ | 1.3 [1.0-1.6] | 1.3 [1.0- 1.5] |
| Percent increase in tx rate, last 20 yrs | $change_{\omega}$ | 0.14 [0.04-0.28] | 0.14 [0.06- 0.27] |
| Treatment success ratio | $k$ | 0.82 [0.67-0.96] | 0.83 [0.69- 0.95] |
| Relative risk of reinfection | $\chi$ | 0.59 [0.08-0.97] | 0.50 [0.24- 0.93] |

Table shows posterior parameter distributions for each of 13 parameters under the Comparator Model and the Clearance Model. Distributions are presented as means with 95% uncertainty intervals in brackets. See SI Text 1 for more details. All rate parameters are presented as annual rates.

**Supplementary Table 6: Modeled infection dynamics, main analysis and sensitivity analyses**

| Analysis | Model | Percent of *Mtb* infection prevalence that is recent (vs. remote) | Percent of TB disease incidence from recent infection (vs. remote) | One-year avertable fraction of TB incidence* | Annual Risk of Infection |
|---|---|---|---|---|---|
| **Main Analysis** | **Comparator** | 1.1% [0.7-2.2%] | 28% [14-46%] | 14% [8-23%] | 1.1% [0.8-1.5%] |
| | **Clearance** | 8.0% [3.6-14.7%] | 61% [35-79%] | 32% [20-44%] | 4.8% [2.5-7.3%] |
| **50% of cleared infections remain immunoreactive** | **Comparator*** | 1.1% [0.7-2.2%] | 28% [14-46%] | 14% [8-23%] | 1.1% [0.8-1.5%] |
| | **Clearance** | 8.6% [3.8-17.0%] | 57% [37-77%] | 30% [17-40%] | 3.4% [2.1-5.8%] |
| **All cleared infections remain immunoreactive** | **Comparator*** | 1.1% [0.7-2.2%] | 28% [14-46%] | 14% [8-23%] | 1.1% [0.8-1.5%] |
| | **Clearance** | 4.8% [0.9-13.3%] | 43% [21-71%] | 23% [11-37%] | 1.6% [1.0-2.6%] |
| **Declining effective contact rate** | **Comparator** | 0.6% [0.3-1.1%] | 16% [8-26%] | 8% [4-13%] | 0.7% [0.4-1.0%] |
| | **Clearance** | 4.4% [1.4-12.0%] | 49% [26-71%] | 27% [14-45%] | 4.8% [2.5-7.3%] |

The "one-year avertable fraction" represents the fraction of new incident disease in a given year that would be averted if all people with TB disease at the start of that year were instantaneously diagnosed and treated at year-start. All results show means across 20,000 model simulations, with corresponding 95% uncertainty intervals in parentheses.

* Since changing the proportion of cleared infections remaining immunoreactive has no effect in a model without clearance, these results are copied over from the top row of the table.

**Supplementary Table 7: Duration of TB disease, main analysis and sensitivity analyses**

| Analysis | Model | Average duration of disease with current treatment patterns (in years) | Average duration of disease absent treatment (in years) |
|---|---|---|---|
| **Main Analysis** | **Comparator** | 1.5 [1.2-1.8] | 2.2 [1.7-2.8] |
| | **Clearance** | 1.5 [1.3-1.8] | 2.1 [1.7-2.8] |
| **50% of cleared infections remain immunoreactive** | **Comparator*** | 1.5 [1.2-1.8] | 2.2 [1.7-2.8] |
| | **Clearance** | 1.6 [1.2-2.0] | 2.2 [1.7-2.9] |
| **All cleared infections remain immunoreactive** | **Comparator*** | 1.5 [1.2-1.8] | 2.2 [1.7-2.8] |
| | **Clearance** | 1.7 [1.3-2.1] | 2.5 [1.7-3.1] |
| **Declining effective contact rate** | **Comparator** | 1.4 [1.2-1.7] | 2.1 [1.7-2.7] |
| | **Clearance** | 1.7 [1.3-2.2] | 2.3 [1.7-3.0] |

Estimates are shown as means in the average duration across 20,000 model simulations, with corresponding 95% uncertainty intervals in parentheses.

* Since changing the proportion of cleared infections remaining immunoreactive has no effect in a model without clearance, these results are copied over from the top row of the table.

**Supplementary Figure 3: Projected impact of case finding and mass TPT provision on incidence**

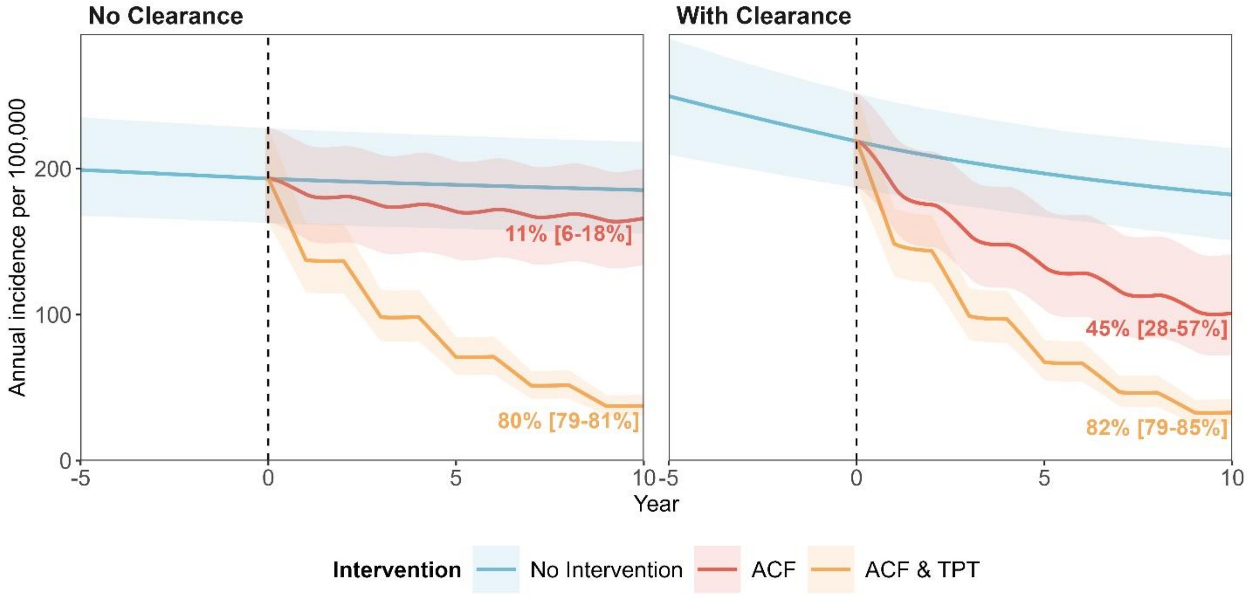

Figure shows the projected TB incidence per 100,000 people over ten years of simulated interventions (years 0 through 10), including: no intervention (blue), Active Case Finding/ACF waves every 3 years (red-orange), adding TPT to the ACF waves (yellow). Panels differ by whether the model structure includes infection clearance (left panel = Comparator Model without clearance; right panel = Clearance Model).

**Supplementary Figure 4: Projected impact of case finding and mass TPT provision on prevalence**

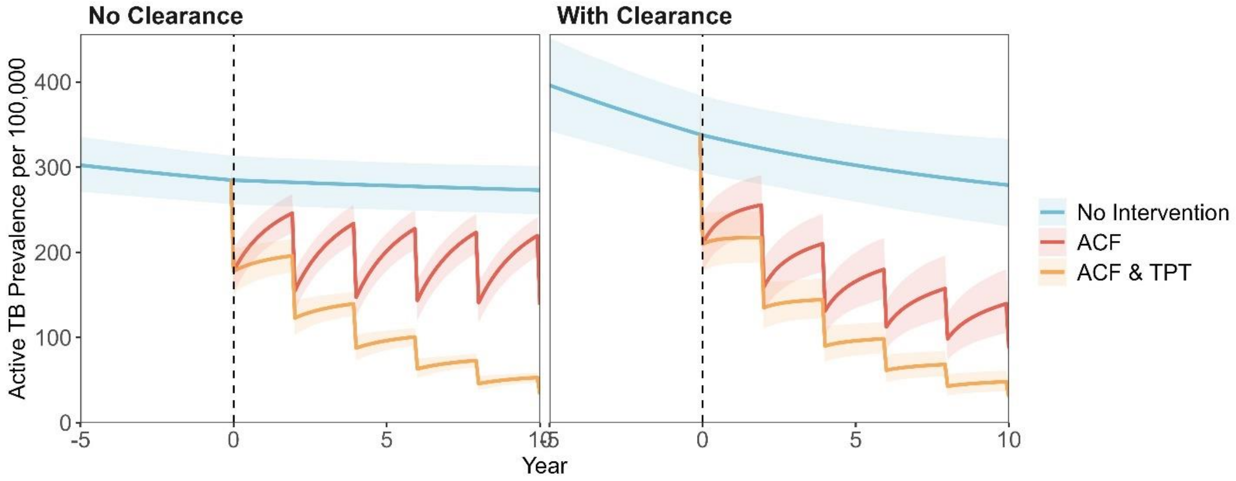

Figure shows the projected prevalence of TB disease per 100,000 people over ten years of simulated interventions (years 0 through 10), including: no intervention (blue), Active Case Finding/ACF waves every 3 years (red-orange), adding TPT to the ACF waves (yellow). Panels differ by whether the model structure includes infection clearance (left panel = Comparator Model without clearance; right panel = Clearance Model).

**Supplementary Figure 5: Projected impact of case finding and mass TPT provision on TB mortality**

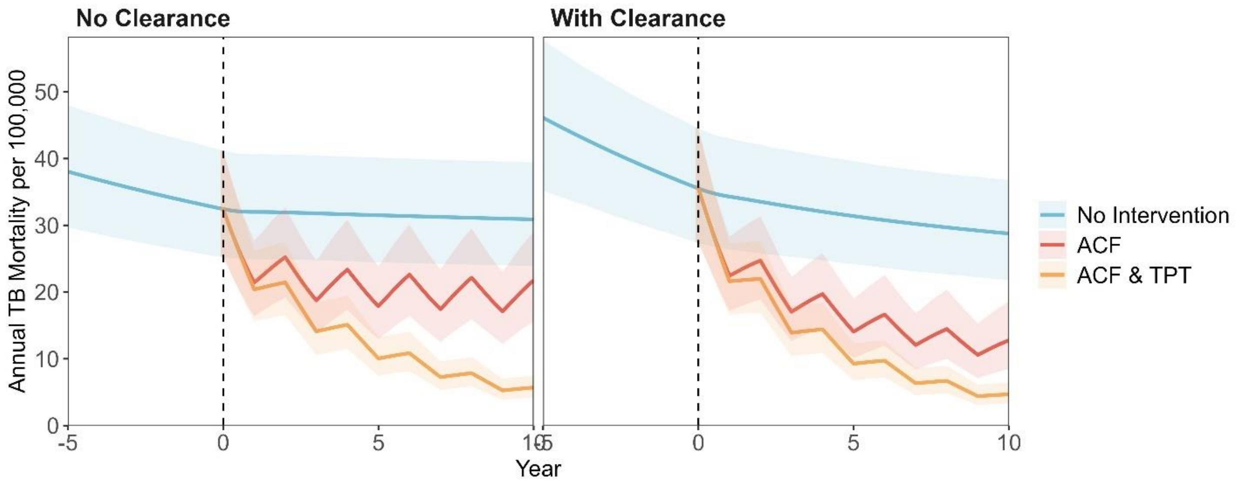

Figure shows the projected annual TB mortality per 100,000 people over ten years of simulated interventions (years 0 through 10), including: no intervention (blue), Active Case Finding/ACF waves every 3 years (red-orange), adding TPT to the ACF waves (yellow). Panels differ by whether the model structure includes infection clearance (left panel = Comparator Model without clearance; right panel = Clearance Model).

**Supplementary Figure 6: Fit of Conventional and Clearance models to 9 calibration targets (sensitivity analysis on % of cleared infections that remain immunoreactive)**

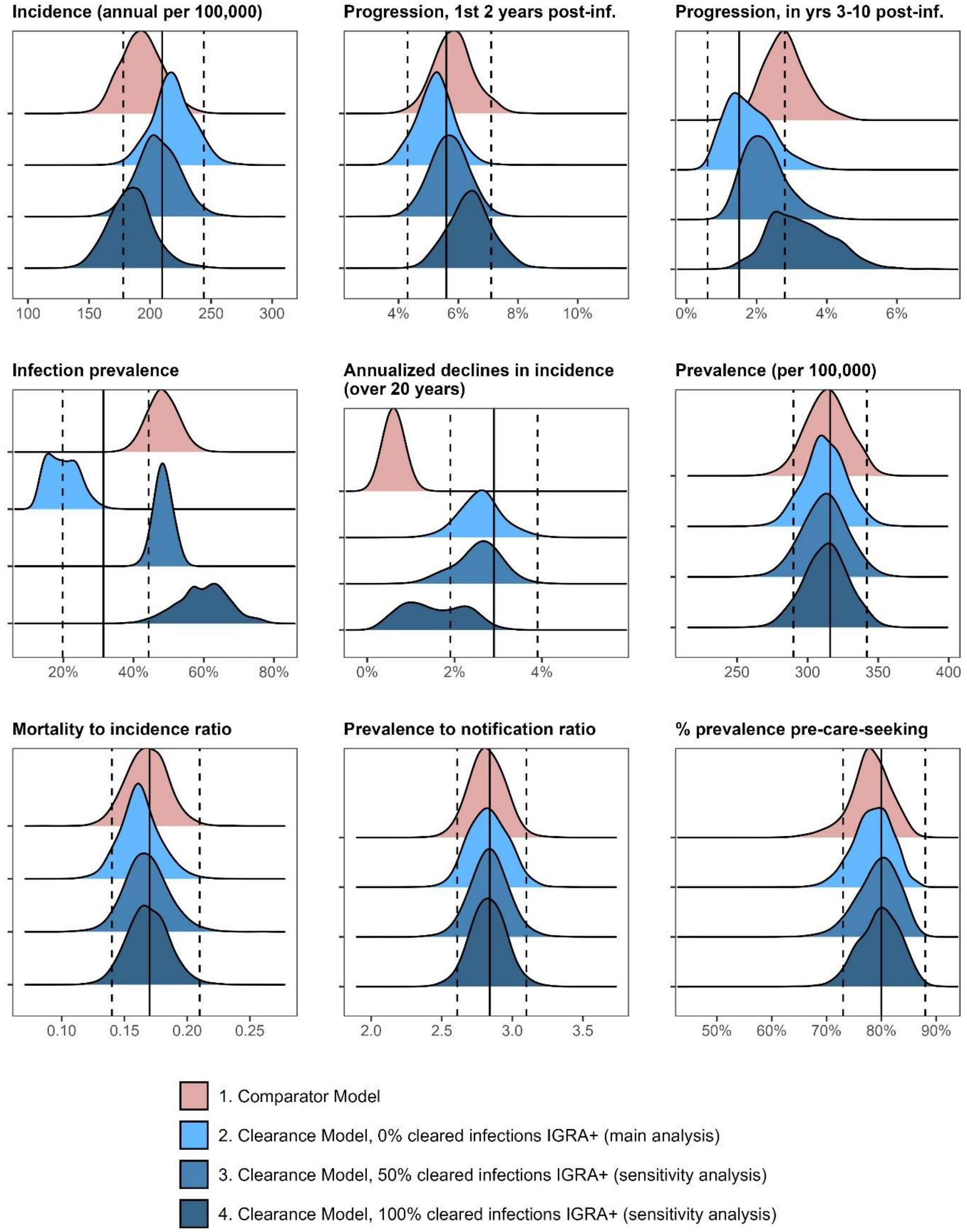

Figure shows the posterior distributions of 9 modeled outputs (shaded density plots) against corresponding calibration targets (mean values indicated by solid black vertical lines, 95% confidence intervals indicated by grey shading with dashed vertical lines. Outputs from the Clearance Model are shown in blue and outputs from the Comparator Model (no clearance) are shown in red. Darker blue distributions (bottom 2 distributions on each panel) indicate sensitivity analyses in which 50% or 100% of cleared infections were assumed to remain immunoreactive/IGRA-positive; the main analysis (0% cleared infections IGRA-positive) is shown in lighter blue.

**Supplementary Figure 7: Fit of Conventional and Clearance models to 9 calibration targets (sensitivity analysis on mechanism inducing pre-intervention declines in incidence)**

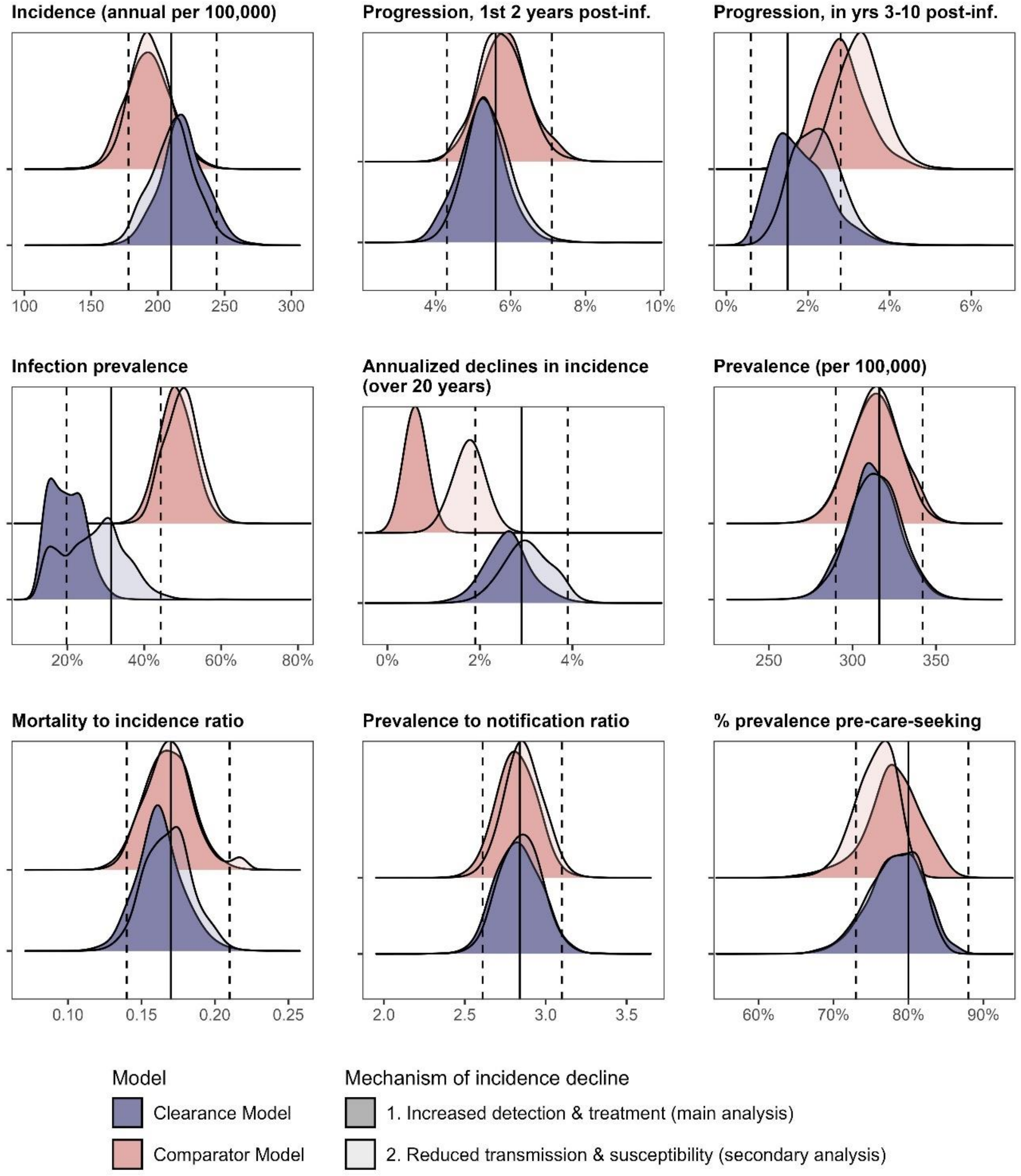

Figure shows the posterior distributions of 9 modeled outputs (shaded density plots) against corresponding calibration targets (mean values indicated by solid black vertical lines, 95% confidence intervals indicated by grey shading with dashed vertical lines. Outputs from the Clearance Model are shown in blue and outputs from the Comparator Model (no clearance) are shown in red. Outputs from the main analysis, in which declines in TB incidence during the pre-intervention period were induced via increases in the treatment initiation rate, are shown in darker shading. Outputs from the secondary analysis, in which declines in TB incidence were instead induced via reductions in the effective contact rate, are shown in lighter shading.

**Supplementary Figure 8: Projected impact of case finding and mass TPT provision on TB incidence (sensitivity analyses on the proportion of cleared infections that maintain immunoreactivity)**

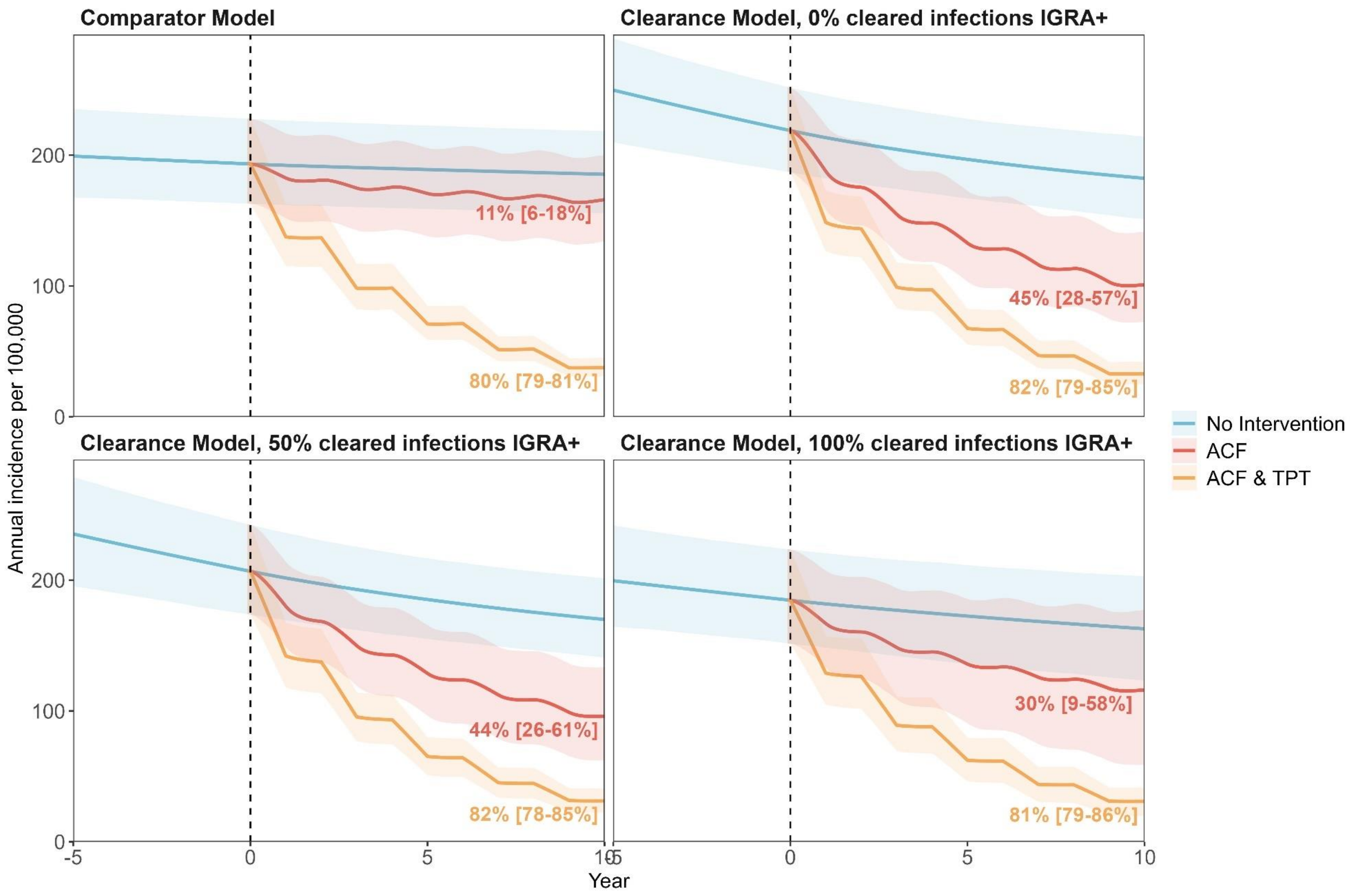

Figure shows the projected annual TB incidence per 100,000 people over ten years of simulated interventions (years 0 through 10), including: no intervention (blue), Active Case Finding/ACF waves every 3 years (red-orange), adding TPT to the ACF waves (yellow). Panels differ by whether the model structure includes infection clearance and, if it does, what was assumed about immunoreactivity among people with cleared infection: top-left panel = Comparator Model without clearance (main analysis); top-right panel = Clearance Model with 0% of cleared infections remaining immunoreactive (main analysis); bottom-left panel = Clearance Model with 50% of cleared infections remaining immunoreactive (sensitivity analysis); bottom-right panel = Clearance Model with 100% of cleared infections remaining immunoreactive (sensitivity analysis);

**Supplementary Figure 9: Projected impact of case finding and mass TPT provision on TB incidence (sensitivity analyses on the mechanism of pre-intervention incidence declines)**

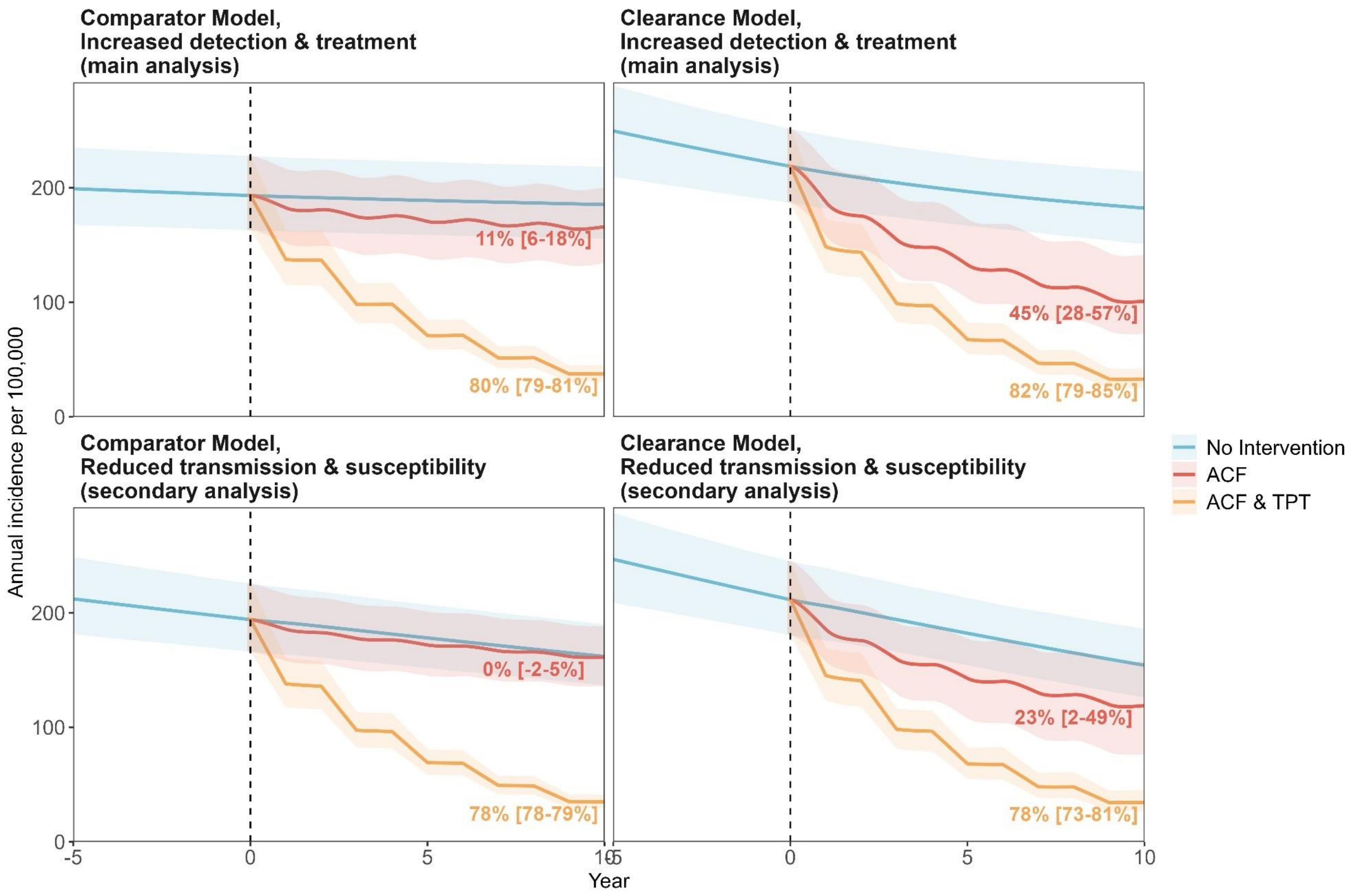

Figure shows the projected annual TB incidence per 100,000 people over ten years of simulated interventions (years 0 through 10), including: no intervention (blue), Active Case Finding/ACF waves every 3 years (red-orange), adding TPT to the ACF waves (yellow). Panels differ by whether the model structure includes infection clearance (left panels = Comparator Model without clearance; right panels = Clearance Model) and the mechanism by which incidence declines were induced in the pre-intervention period (top panels = main analysis, via increases in the detection and treatment rate; bottom panels = sensitivity analysis, via reductions in the effective contact rate).

**Supplementary Figure 10: Impact of ACF at varying ACF and TPT coverage and effectiveness**

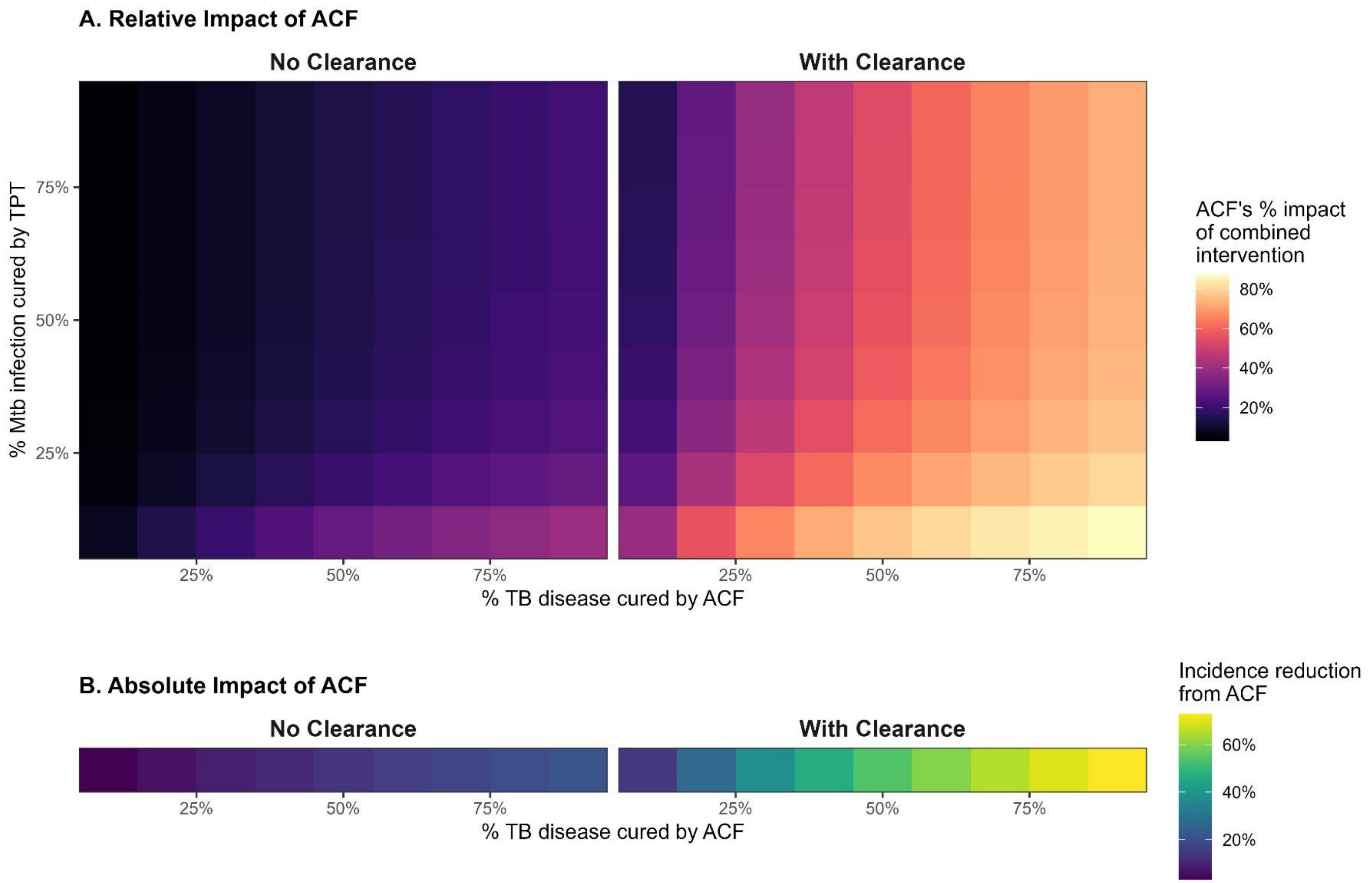

Panel A shows the % of incidence reduction at 10 years from the combined intervention of active case-finding (ACF) and TPT that is attributable to ACF alone over varying levels of ACF coverage/efficacy (x-axis; % of active TB disease that is cured by each intervention wave) and TPT coverage/efficacy (y-axis; % *of Mtb* infection that is cured by each intervention wave). Panel B shows the incidence reduction at 10 years from ACF alone, over varying ACF coverage/efficacy (same x-axis as Panel A). Sub-panels differ by whether the model includes infection clearance (left = Comparator Model without clearance; right = Clearance Model).

**Supplementary Figure 11: One-way sensitivity analysis on impact of ACF on TB incidence**

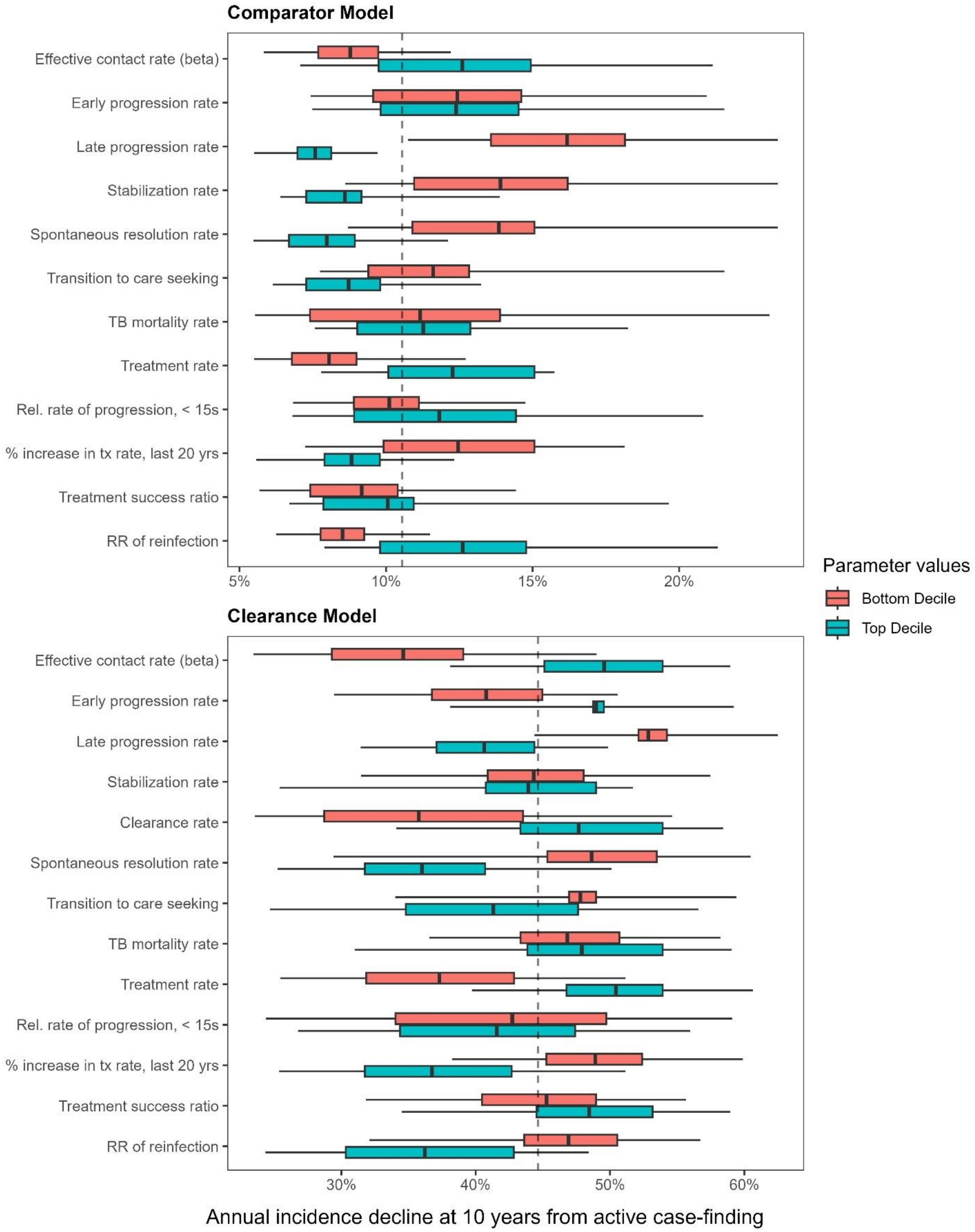

Figure shows how the decline in incidence at 10 years from active case-finding/ACF varies across the range of each model parameter. In each panel, each pair of boxplots shows variation in this outcome (incidence decline at 10 years from active case-finding) when the analysis was limited to either simulations in which the value of the parameter of interest was in the top (green-blue) or bottom (red) decile of its values across all posterior parameter sets. In each boxplot, the edges of the box represent the interquartile range, the band in the middle represents the mean, and the end of the whiskers represent 2.5$^{th}$ and 97.5$^{th}$ percentiles. Vertical dotted line show means across all posterior parameter sets samples.

**Supplementary Figure 12: Correlations between intervention impact & calibrated model parameters**

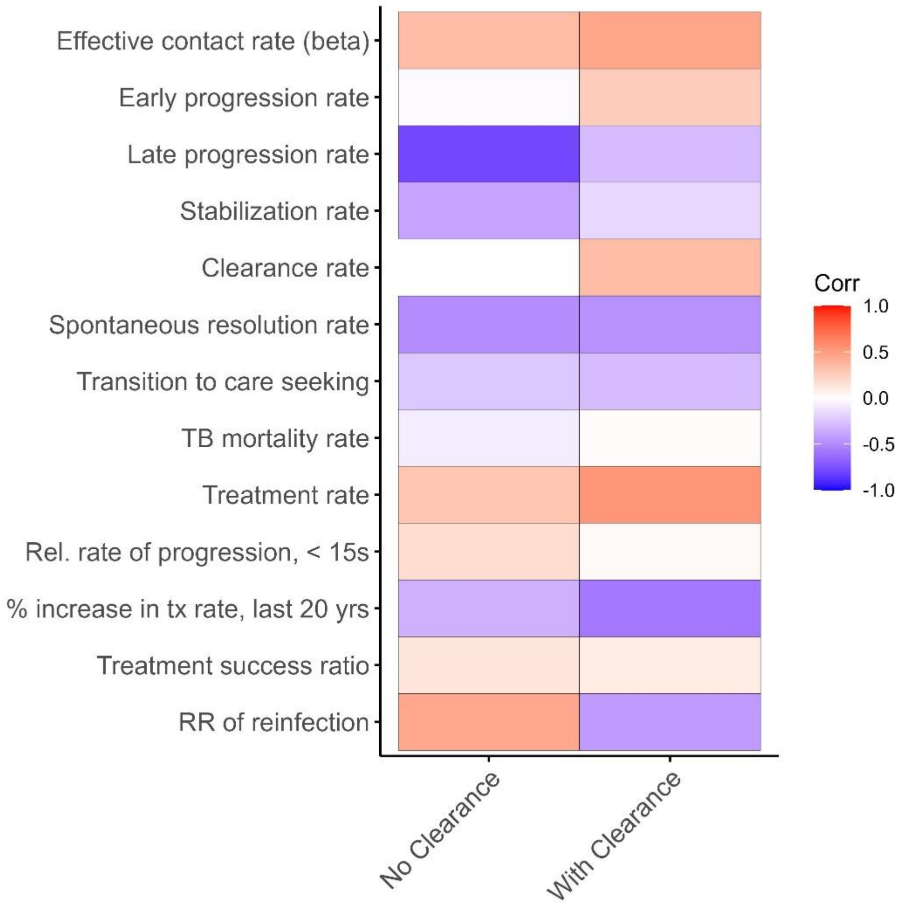

Figure shows pairwise correlations between model parameters (vertical axis) and intervention impact (incidence declines at 10 years with active case-finding), under both model variations (horizonal axis).
